# Brain network and blood transcriptomic correlations underpin psychopathological phenotypes: A Preliminary Study

**DOI:** 10.1101/2025.07.08.25331132

**Authors:** Dennis Wylie, Dhivya Arasappan, Uttam Khatri, Madeleine Dwortz, Mbemba Jabbi

**Affiliations:** Center for Biomedical Research Support, The University of Texas at Austin, USA; Department of Psychiatry and Behavioral Sciences, The University of Texas at Austin Dell Medical School, Austin, Texas, USA; Center for Learning and Memory, The University of Texas at Austin, Dell Medical School, Austin, Texas, USA; Mulva clinics for the Neurosciences, Dell Medical School, Austin, Texas, USA

## Abstract

Depressive disorders and comorbid psychopathologies affect over 12% of humans and present debilitating symptoms. Prior studies identified systemic brain and peripheral biological dysregulations in depressive disorders, but the interrelated brain-body molecular pathobiology underlying these conditions is not well-defined. Here, we studied the molecular signatures of depressive and comorbid psychopathology in the same 14 postmortem brain anterior insula (Ant/Ins) and subgenual cingulate (sgACC) tissue and whole blood donors. We showed that the brain Ant/Ins and sgACC regions exhibited positive (concordant) brain-brain correlations in ∼50% and less than 10% negative (discordant) brain-brain correlations in our observed gene expression markers. In contrast, our observed gene expression/transcriptomic correlations between the brain regions and blood (brain-blood correlations) were more normally distributed, such that ∼20% were concordantly, and ∼20% were discordantly correlated. Using a mixed effects analysis, we showed that increased measures of comorbid psychopathology were associated with selective upregulation of inflammatory cytokine receptor gene *TEC* and neurodevelopmental olfactory receptor genes *OR52E4* and *OR56B2P* transcriptomes across the two brain regions and blood. In contrast, an upregulation of the proinflammatory cytokine receptor gene *IL18R1* and cellular developmental gene *WIF1* and a downregulation of the brain protein signaling gene *TECTB* were more associated with comorbid psychopathology in blood than in the brain. Our findings of concordant and discordant transcriptomes in a well-phenotyped sample of 14 multi-tissue donors revealed tightly correlated brain-blood transcriptomic underpinnings of comorbid psychopathology. Our results provide a framework for identifying targetable brain-derived peripheral markers to advance novel diagnostic and therapeutic development for psychopathologic conditions.

## INTRODUCTION

Neuropsychiatric disorders such as major depressive disorder (MDD) often co-occur with other conditions and present debilitating psychopathological symptoms leading to significant disease burden (Kessler et al. 2009; Stein et al. 2022; McGrath et al. 2023; Solmi et al. 2023). Despite the high prevalence of MDD and comorbid conditions, the brain and peripheral molecular dysfunctions underlying psychopathology are not well-defined (Glannon, 2022; Abi-Dargham et al. 2023; Doerig et al. 2023). This lack of understanding of the link between brain and peripheral molecular markers underlying severe psychopathology is, in part, due to a lack of access to human brain tissue in vivo. If peripheral biomarkers could be systematically linked to brain correlates for psychopathology, novel diagnostic and therapeutic interventions could be developed.

Parallel studies targeting blood (Bhagar et al. 2024) and brain (Arasappan et al. 2025) molecular measures associated with psychopathology have contributed novel insights. However, efforts to examine the direct relationship between brain and blood molecular markers of psychopathology in the same study samples are limited. A recent human study of MDD suicide, MDD non-suicide, and controls with no psychopathology (Mamdani et al. 2022), as well as a preclinical study of mouse models of alcohol dependence (Ferguson et al. 2022) both examined brain and blood samples and found correlated brain – blood transcriptomic signatures for pathobiological phenotypes. While the human study included 45 postmortem donors with both brain and blood samples, the blood RNA quality as measured with RNA integrity numbers included very low-quality blood samples, making it challenging to validate the biologically conserved nature of the observed brain to blood molecular correlations (Mamdani et al. 2022).

Of relevance to studies of brain–blood molecular overlaps, preclinical studies have shown that myeloid (bone marrow) derived blood cells such as monocytes can cross the blood-brain barrier (BBB) and differentiate into glia and in less frequency to neurons in brain injury phenotypes (Mezey et al. 2005). Further evidence supports the existence of mechanisms by which myeloid-derived peripheral blood mononuclear cells (PBMCs) or hematopoietic stem cells (HSCs) transdifferentiate into brain glial and neuronal cellular lineages during increased disease phenotypes (Mezey et al. 2000; Corti et al. 2002; Hodes et al. 2015) such as stress-related phenotypes like MDD are causally linked to pathological brain-blood cellular and molecular interactions (Poller et al. 2022; Cathomas et al. 2024). Of interest, cellular pathologies such as higher monocyte counts and related dysregulated interleukin 6 (*IL6)* signaling in the periphery underpins increased psychopathological phenotypes (Schiweck et al. 2020; Serna-Rodriguez et al. 2022; Bai et al. 2023); suggesting the existence of cellular and molecular crosstalk between brain and peripheral blood. Whether such brain–blood molecular interactions may underpin depressive disorder and comorbid phenotypes, postmortem remains underexplored.

Here, we studied the transcriptomic signatures in the brain anterior insula (Ant/Ins) and subgenual anterior cingulate (sgACC) in light of these brain regions critical regulatory roles in coding bodily feeling states like pain, itch, exteroceptive temperature sensation, and abstract mood and emotion states (Jabbi et al. 2008; Harrison et al. 2009; Joyce et al. 2018; Levinthal et al. 2020). We applied deep RNA-seq at 120 million reads/sample in bulk brain tissue dissections from the Ant-Ins and sgACC, as well as whole blood separately. The study aims to identify overall correlation patterns between the brain regions and blood and assess the significance of these correlations in the context of psychopathological phenotypes (Egervari 2019; Craske et al. 2023; Sun et al. 2024). We tested the hypothesis that the translational readout of brain network molecular signatures will tightly correlate with blood molecular signatures. This correlation would allow us to select RNA expression signals represented in brain and blood (biomarkers) related to psychopathological phenotypes.

### Concordant and Discordant Molecular Pathological Signatures in Brain & Blood

We first examined the degree of global correlation and directionality (positive ‘concordant’ or negative ‘discordant’) of our observed global correlations between the calculated mean RNA read counts of gene expression markers identified in a) Ant/Ins brain region and b) sgACC brain region (brain – brain correlation). Most importantly, we further assessed the global correlations between the Ant/Ins and blood (brain–blood correlation) and between sgACC and blood (brain–blood correlation). To this aim, we performed the Spearman rank correlation analysis of the global molecular/gene expression marker expression signatures across the entire Rho range of -1 to +1, including protein-coding genes, characterized and uncharacterized transcripts, non-coding RNA variants, and isoforms/spliced variants. We then applied the Kolmogorov-Smirnoff test to identify the distribution patterns of our observed brain–brain and brain–blood correlations. This analysis allowed the mapping out of the global distribution of the identified correlations between the two brain regions relative to each brain region and blood. It enabled the generation of novel hypotheses regarding the molecular signatures underpinning psychopathology in the brain and blood.

We found that in the Ant/Ins and sgACC brain–brain correlations, 48.34% of markers correlated concordantly, whilst only 3.12% of markers correlated discordantly, in addition to 43.34% of markers showing no global brain–brain correlations (**Fig 1 A-C**). By assessing the Ant/Ins – blood (brain–blood) correlations, we found that 21% of the markers correlated concordantly, whilst 17.78% of markers correlated discordant, and a relatively higher (compared with the brain–brain correlated) distribution at 61% of markers showed no Ant/Ins – blood correlation (**Fig A**, and **C**). Similarly, our observed global correlations between brain ‘sgACC’ – blood was concordant in 17.68% of markers and discordant in 16.36% of markers, whereas 65.95% of markers showed no brain ‘sgACC’ – blood correlations (**Fig 1 B** and **C**). As expected, the comparisons of the correlation distributions between Ant/Ins – blood and sgACC– blood were not significantly different (Fig. 1 C). In contrast, the comparisons of the global correlation distributions between brain – brain, relative to the Ant/Ins – blood ‘brain-blood’ and sgACC – blood ‘brain-blood’ correlations were not similarly distributed (Fig. 1 A and B).

**Figure 1.**
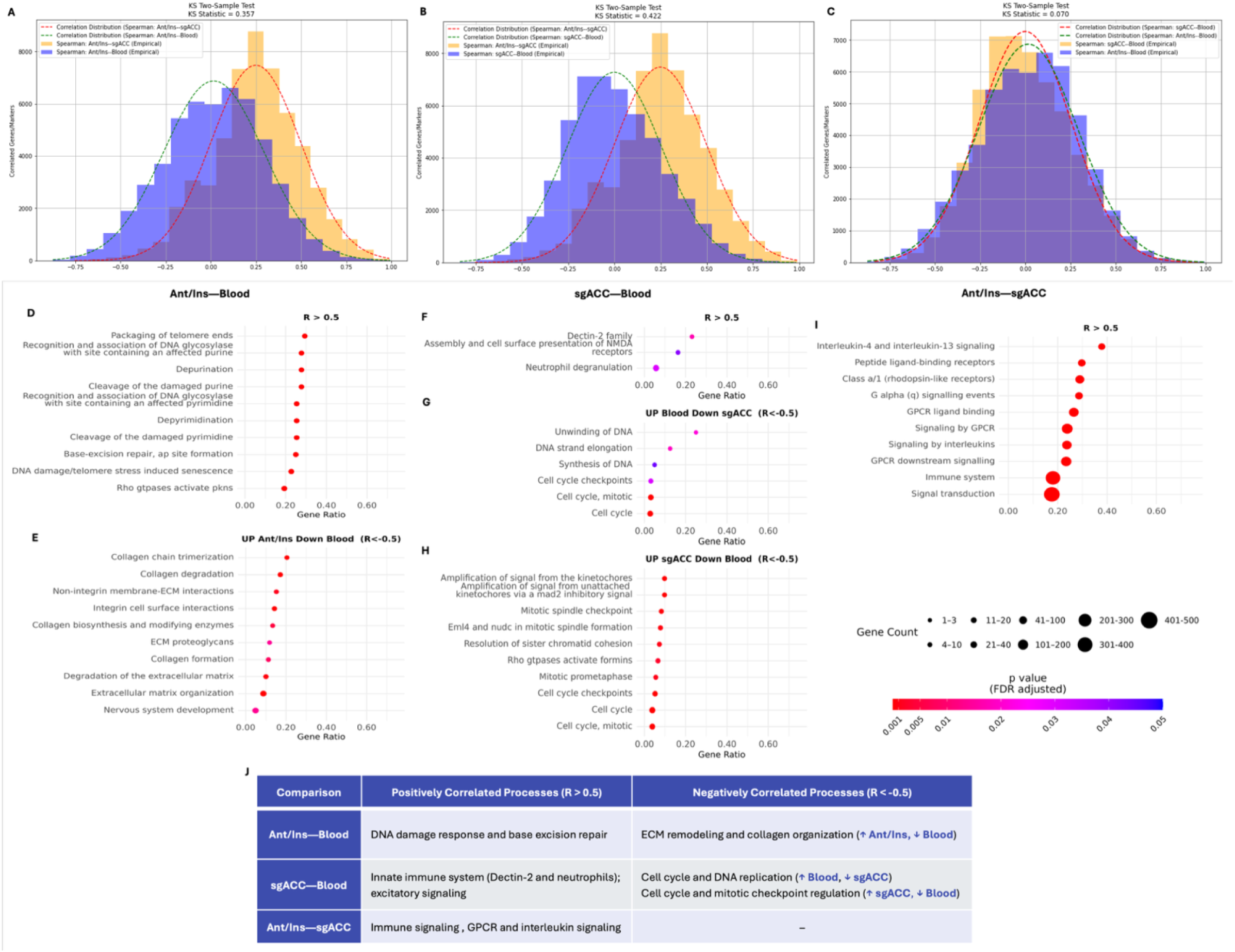
Concordant and Discordant Transcriptome Correlations between the read counts identified in the Ant/Ins and sgACC, Ant/Ins – Blood, and sgACC – Blood. A-C illustrates transcriptomic markers (including protein coding, non-protein coding, uncharacterized genes/RNA, and spliced variants) distribution across brain and blood RNA-seq readcounts. A show the differences in read count distribution between Ant/Ins – sgACC (brain – brain) correlations compared with Ant/Ins – blood (brain – blood) correlations using the Kolmogorov Smirnov (KS) 2-sample test with effect size of 0.357 in correlation distribution difference. B shows a similar KS statistical difference between Ant/Ins – sgACC correlation compared with sgACC – blood correlation with KS 2-sample effect size of 0.422. C illustrates no differences between the distribution of Ant/Ins – blood correlation compared with sgACC – blood correlation with KS stats effect size of 0.07. (D–J) Reactome pathway enrichment analysis of genes with positive (R > 0.5) or negative (R < –0.5) correlations between brain regions and blood is shown in panels D–I. Genes positively correlated between Ant/Ins and blood are enriched for DNA damage recognition and base excision repair pathways (D). Negatively correlated genes with higher expression in Ant/Ins are enriched for extracellular matrix remodeling and collagen-related processes (E). Genes positively correlated between sgACC, and blood are enriched for innate immune signaling (F). Negatively correlated genes with higher expression in blood are enriched for DNA replication and cell cycle processes, while those with higher expression in sgACC are enriched for mitotic checkpoint and spindle-related functions (G, H). Genes positively correlated between Ant/Ins and sgACC are enriched for immune and GPCR signaling pathways (I). A summary of the pathways associated with positively and negatively correlated gene sets across each comparison is shown in (J).

We then analyzed gene ontology to assess the functional genetic pathways underlying the observed concordant and discordant correlations. The top 5% (i.e., biologically meaningful correlations ranging between >= 0.75 concordant correlations and <=-0.75 discordant correlations between the Ant/Ins – sgACC and Ant/Ins – blood as well as sgACC – blood for the same protein-coding and non-protein-coding markers. We found predominant cell cycle and cellular developmental signaling pathway enrichments (**Fig 1 D-I**) associated with brain-blood correlated markers. These global correlation patterns suggest that the proportion of convergently expressed genes/transcriptomes is similar to that of discordantly expressed transcriptomes (discordant correlations) in the brain and blood. The observed concordant and discordant brain–blood correlation distribution associated with the cell cycle was not found between the brain regions.

### Psychopathology-related molecular signatures across brain and blood

We applied mixed effects analysis to assess the main effects of the degree of psychopathology/psychiatric morbidity and the potential interaction between the degree of psychopathology and tissue type in types of brain versus blood gene expression using the statistical threshold of p<0.005 (uncorrected) as significant levels. To this aim, we used the number of lifetime comorbid mental health diagnoses (i.e., depression, bipolar disorder, schizophrenia, and records of lifetime mania, anxiety, psychosis, etc., and metrics of the respective severity of these symptoms (i.e., recurrent depression) in the postmortem metadata, to calculate the total of the degree of psychopathology/mental health diagnostic comorbidity coupled with co-occurring symptom complexity in the same individual donor. For this measure, we created two comparison groups of *high* (with scores of degrees of psychopathology from 4 to 7 comorbid diagnoses/symptoms) versus *low* (with scores of degrees of psychopathology from 0 to 3 comorbid diagnoses/symptoms).

By adjusting for age, sex, and RNA integrity numbers (RINs), our mixed-effects analysis revealed a main effect of high versus low psychopathology/morbidity in all three tissue types (i.e., Ant/Ins brain, sgACC brain, and blood) was associated with protein tyrosine kinase marker *TEC* involved in the intracellular signaling mechanisms of cytokine receptors, lymphocyte surface antigens, heterotrimeric G-protein coupled receptors, and integrin molecules, in addition to two neurodevelopmentally critical olfactory receptor genes *OR52E4* and *OR56B2P* (**Fig 2**). We further found the main effects of high versus low psychopathology/morbidity in all three tissue types (i.e., Ant/Ins brain, sgACC brain, and blood) in the expression of non-protein coding markers LINC02843 and UNC93B2.

**Figure 2.**
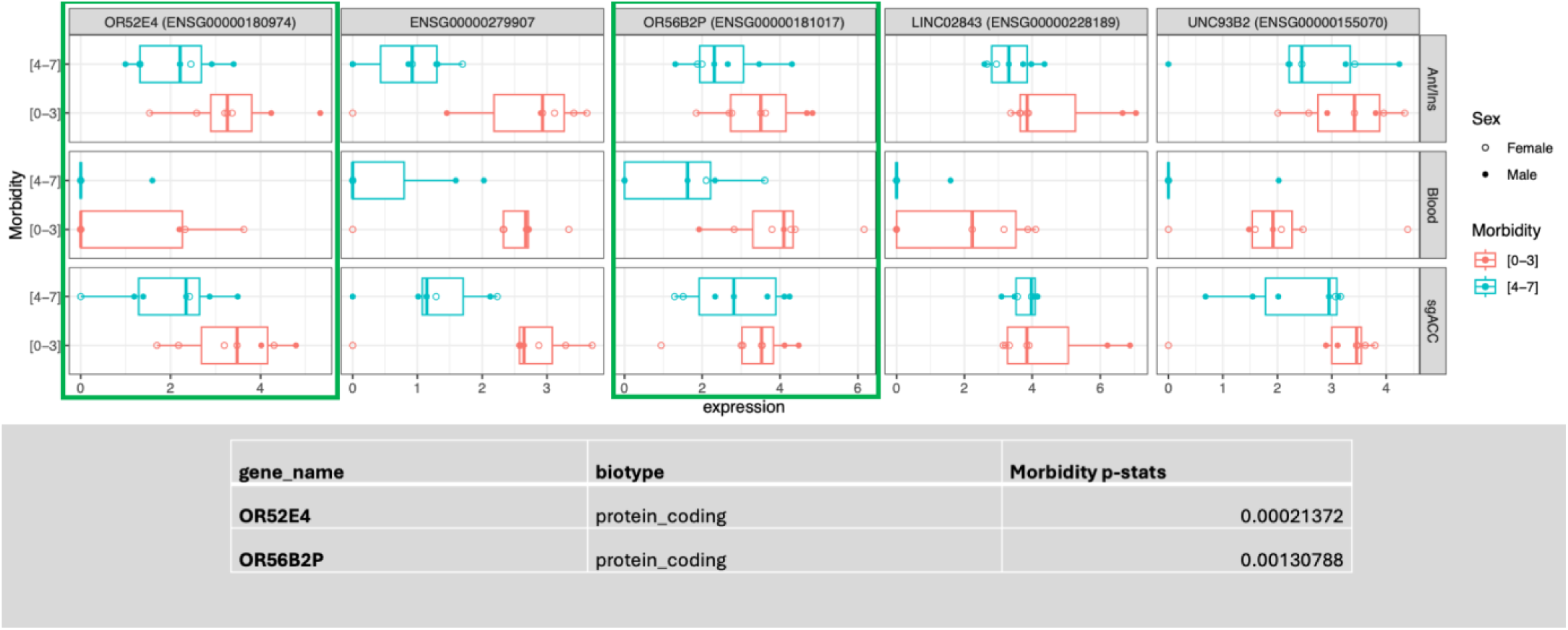
Main effect of comorbid psychopathology. The figure illustrates the total number of observed transcriptome markers (5 in total) including two olfactory receptor protein encoding genes that were differentially expressed similarly in the two brain regions and blood in related to the degree of comorbid lifetime psychopathology.

In addition to our observed main effects of psychopathology/mental illness morbidity or complexity across the 14 donor samples independent of tissue type, we also assessed the interaction between biological tissue type (Ant/Ins, sgACC, and blood) and increased psychopathology/morbidity (i.e., high > low psychopathology/morbidity). For this mixed effect interaction analysis, we tested the hypothesis that the interaction between psychopathology/morbidity and tissue type would reveal greater anomalies in central nervous system (CNS)-related gene expression marker enrichment compared to anomalies in peripheral blood inflammatory markers associated with psychopathology. In line with our hypothesis, we found an interaction between increased degree of psychopathology (high versus low) BY sample type (Ant/Ins, sgACC, and blood) such that increased psychopathology was associated with more downregulation of 5 genes *TECTB*, *WIF1*, *GBA3*, *EPS8L3*, and *DMRT3* in blood than the two brain regions. In contrast, we further found that four genes, *IL18R1*, *EIF1AY*, *IP6K3*, and *NPIPB15,* were significantly dysregulated (i.e., downregulated or upregulated) along with 16 long non-protein coding RNAs/pseudogenes in blood than in the two brain regions in individuals with high vs. low psychopathology/morbidity suggestive of blood associated psychopathological markers not enriched in the studied brain regions (**Supplementary Fig 1**). These observed correlated markers were related to immune/inflammatory gene expression signatures markedly present in blood compared with the brain regions in donors with increased comorbid psychopathology (**Supplementary Fig 2**).

Given that **80%** of our observed transcriptome expression markers correlated in the two brain regions, we further conducted a direct comparison of blood versus brain transcriptome to identify peripheral biomarkers that might not have strong brain correlates in relation to the degree of psychopathology. We found 18 predominantly immune/inflammatory genes, of which 11 were more upregulated in the differential expression analysis at p<0.005 in blood *LILRB3*, *IL18R1*, *CCL3*, *CCL2*, *CSF3*, *CH25H*, *TNFRSF17*, *IL1RL1*, *CCL3L1*, IL1R2, and PCK1 in donors with higher degrees of lifetime psychopathology, coupled with 7 protein-coding genes were found to be more downregulated in at p<0.005 in blood than brain *CCL8*, *CXCR1*, *SLC11A1*, *CXCL8*, *MT1X*, *HAMP*, and *IL6* in donors with increased psychopathology (**Supplementary Fig 2**).

### Brain and Blood Molecular Signatures for Depressive Disorder

To assess the more diagnostically proximate significance of our observed brain–blood transcriptomic correlations within the context of how the concordant and discordant correlations are associated with lifetime depressive disorder, we conducted differential gene expression analyses differences in donors with lifetime depressive disorder (n=9) versus non-depressed donors (n=6). We then applied a Spearman correlation analysis of brain–brain and brain–blood correlations of the differentially expressed genes (DEGs) to identify correlated DEGs.

We included DEGs surviving Log2Foldchanges >=1.5 in our Spearman correlation model between Ant/Ins, sgACC, and blood to assess the depression-related DEG correlations. We set the significant cut-offs using a two-prong approach: only DEGs >= Log2Foldchanges of 1.5 exhibiting Spearman’s Rho ranging between +0.25 to +1.0 (for concordantly correlated markers) and Spearman’s Rho ranging between -0.25 to -1.0 (for discordantly correlated markers). We found 20 markers showing concordant correlations between brain and blood and 11 markers with discordant correlations between brain and blood (**Fig 3 A-F**).

**Figure 3.**
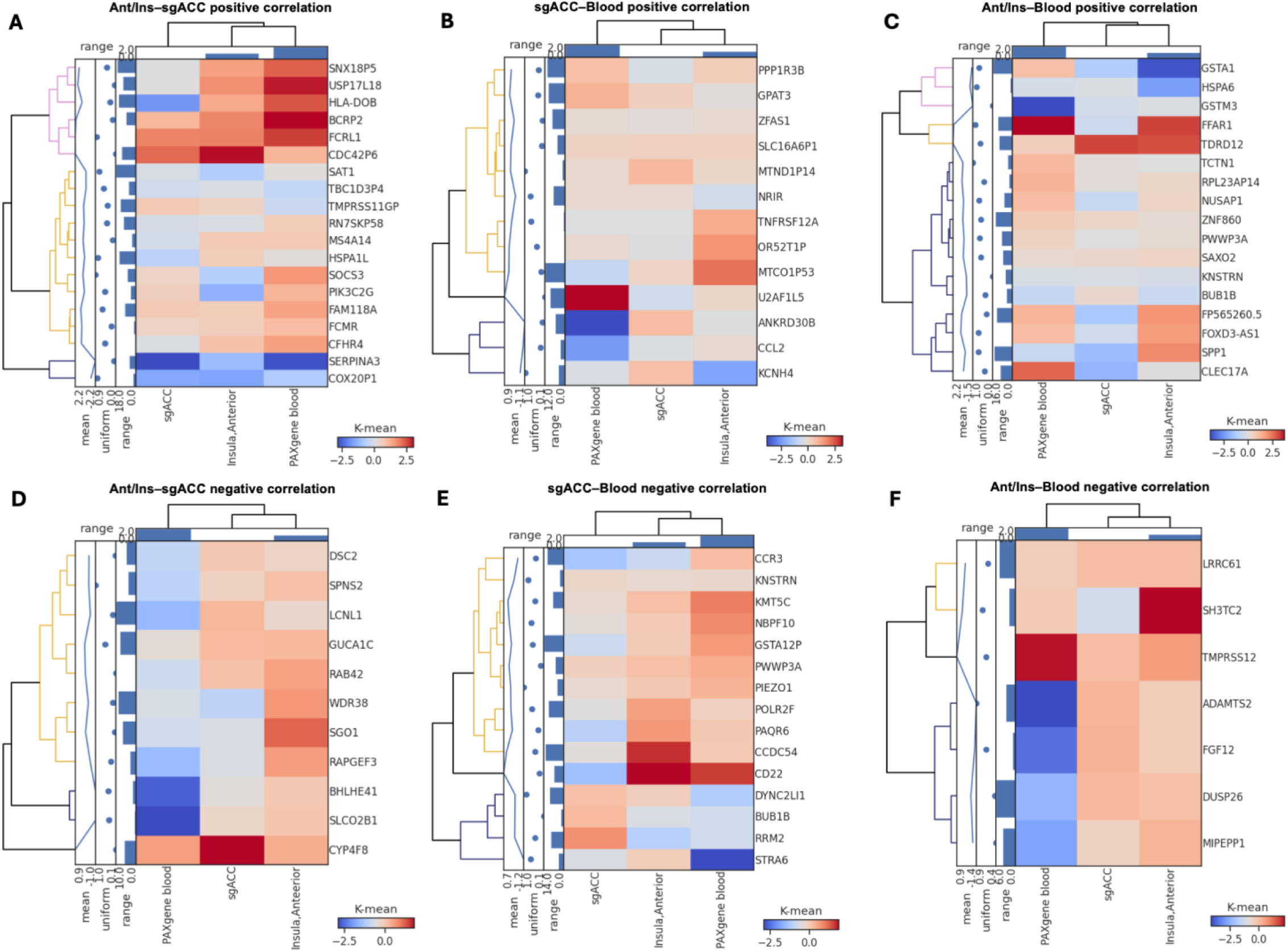
K-means clustering maps of the correlations between brain (sgACC and Ant-Ins) and blood, and related statistics. The shared correlated list of protein coding genes/transcriptome markers, and integrated gene expression levels (in log2fold change values) and correlation scatter plots in Figure 3 illustrates that each of the lists of concordantly correlated or discordantly correlated genes between each of the two studied brain regions and blood in the same sample of 14 donors reflecting Spearman Rank correlations of 0.25 as a cut-off and the genes must be significantly expressed differentially between depressive disorder (MDD and bipolar or depression NOS) versus (>) not depressed at p<0.05 to be considered for the correlation analyses. Heatmap of K-means clustering of differentially expressed genes (DEGs) between the sgACC -- Ant-Ins and blood for depressed vs. non-depressed postmortem donor samples. K-means clustering was applied to these group-related correlation to generate a heatmap for differentially expressed protein-coding genes. The colors on the map represent expression levels: blue signifies the lowest transcriptomic/gene expression and red indicates the highest transcriptomic/gene expression. **A)** shows the concordant (shared) Spearman correlations between sgACC and Ant-Ins (blood included), with additional K-means clustering values of the range, scatterplot/uniformity, and means of the transcriptome expression statistics of each correlated marker/gene). **B)** shows the concordant (shared) Spearman correlations between sgACC and blood (Ant-Ins included), with additional K-means clustering values of the range, scatterplot/uniformity, and means of the transcriptome expression statistics of each correlated marker/gene). **C)** illustrates the concordant (shared) Spearman correlations between Ant-Ins and blood (sgACC included), with additional K-means clustering values of the range, scatterplot/uniformity, and means of the transcriptome expression statistics of each correlated marker/gene). **D, E and F** illustrates the discordant Spearman correlations between sgACC and Ant-Ins (blood included), **D**, sgACC and blood (Ant-Ins included) **E**, and Ant-Ins and blood (sgACC included); with additional K-means clustering values of the range, scatterplot/uniformity, and means of the transcriptome expression statistics of each correlated marker/gene).

## DISCUSSION

In this study, we used the same 14 postmortem donors (42 samples in total). We applied deep RNA-sequencing to investigate whether molecular signatures in brain Ant-Ins (n=14) and sgACC (n=14) network will correlate with blood (n=14) biomarkers associated with psychopathological phenotypes. In addition to our findings of globally concordant and discordant transcriptomic correlates between brain and blood, our analysis revealed generalized brain–blood expressed and biased blood-expressed molecular pathological markers underlying increased comorbid psychopathology and depressive disorder diagnosis. Given that over ∼60% of individuals with a neuropsychiatric diagnosis experience multiple symptoms or comorbid conditions (Kessler et al. 2009), our findings of gene sets showing concordant and discordant expression levels in both brain and blood samples in association with increased lifetime psychopathological phenotypes underscores the feasibility of translating brain molecular pathological markers into disease-associated blood biomarkers.

Specifically, we identified concordant brain and blood transcriptomic signatures associated with increased psychopathology, particularly involving cell cycle and neurodevelopment/olfactory receptor signaling genes. We further uncovered concordant transcriptomes in the brain and blood related to depressive disorders involving cellular metabolic and developmental, DNA damage and repair, and transcriptional regulatory pathway signaling genes. Our findings of discordant transcriptomic correlations between brain and blood were associated with notch signaling and cell cycle pathways. These findings support our hypothesis that concordant and discordant transcriptomic signatures in the brain and blood will underpin molecular psychopathological phenotypes. Whether our findings represent the interactive brain and peripheral cellular homeostatic processes whereby myeloid-derived monocytes and related lineages cross the blood-brain barrier and transdifferentiate into brain cells in depressive/stress phenotypes (Mezey et al. 2000; Cathomas et al. 2024) needs to be replicated with further investigation in larger samples sizes and validated/translated in preclinical models.

Our findings of biased patterns of gene expression signatures in blood, rather than brain network, especially involving immune and inflammatory molecular pathological markers in increased psychopathological and depressive phenotypes, highlights the potential relevance of resident PBMCs and related functional immune and inflammatory molecular processes in homeostatic maintenance and repair of compromised/damaged brain cells in psychopathogy.

The parallels between brain-derived molecular markers in peripheral blood biomarkers are fascinating. The findings of convergence in pathological markers in brain and blood samples of the same postmortem donors suggest the potential utility and feasibility of translating brain markers into blood-based biomarkers. Such brain-derived blood biomarkers could be targeted for developing novel diagnostic and therapeutics for psychopathological conditions, offering a less invasive and more accessible avenue for clinical intervention advances. RNA’s dynamic nature, specificity, sensitivity, therapeutic potential, and ability to be analyzed in less-invasive samples such as whole blood and at high throughput make it a superior biological marker in many brain-related diagnostic and therapeutic contexts with potential peripheral blood correlates (Morris & Mattick 2014; Ferguson et al. 2022). As such, our results of convergent RNA-seq measures of gene expression signatures underlying psychopathological phenotypes in a critical brain network and whole blood samples of the same postmortem donors offer a level of molecular details that DNA, proteins, or metabolites studies (Xu et al. 2022) might not be able to provide. Overall, these correlations between significant DEGs related to depressive disorders in the same individual donors are a powerful illustration of the tight correlations between our observed *brain* molecular pathological markers, on the one hand, with our observed *blood* molecular pathological markers than previously thought.

In light of the observed strengths of our brain-blood correlations both at the general and pathologically defined levels, the study has some limitations: the preliminary nature of the study, coupled with the limited sample size that dampens statistical power beyond preliminary analyses and postmortem context that limits the generalization of our findings by not allowing a direct bench to bedside validation of these findings, are limitations worth noting. Despite these limitations, the demonstration of the feasibility of translating postmortem brain network molecular pathologies into less invasive peripheral blood molecular biomarkers associated with complex behavioral disorders has several translational implications. Our observed results, especially when translated into clinical populations, provide a framework for studying the multisystemic molecular pathological principles that will guide the advancement of novel diagnostic and therapeutic developments for objective characterization of and therapeutics for prevalent psychopathologies.

## METHODS

### Study population, diagnostic determination, and measures of high vs. low psychopathology

Fourteen donor samples (7 females) were included in this study. The donors had a lifetime history of depression, bipolar disorder, and psychosis, as well as controls, age = 40±23, 50% males, PMI=27±8, 8% Asian, 23% African, 69% Caucasian. Specifically, 7 of the donors had a history of lifetime major depressive disorder (MDD), whilst 2 had a lifetime history of depression not otherwise specified (DepNos), 1 had a lifetime history of bipolar disorder with the most recent episode being a depression before death, 2 had a history of substance use disorder, 1 had a history of anxiety, and 1 had no lifetime history of psychiatric disorder. Of the 14 donors, 5 with a depressive disorder died by suicide, and 9 died of natural deaths. 13 donors had a lifetime history of chronic diseases ranging from cardiovascular to metabolic or liver diseases, and one donor had no history of any chronic disease.

### Study Design & Data Analysis

#### Spearman Correlation Analysis

For each of the three pairs of sample types--Ant/Ins vs. sgACC, Ant/Ins vs. blood, and sgACC vs. blood--and for each gene g, we calculated the Spearman correlation coefficient of the 14 normalized data values from the first sample type (e.g., Ant/Ins) with the 14 corresponding normalized data values from the second sample type (e.g., sgACC).

We first focused on the normalized RNA/marker read counts to examine the global distribution of the Spearman correlation values between the two brain regions and blood (brain-blood correlations) and between the two brain regions (brain-brain correlation). We compared the whole distribution range (from -1 to +1 Spearman Rho values) of the readcounts correlations for the brain-blood and brain-brain correlations (**Fig 1A-C**). All markers with read count values of 0, 1, or missing read counts in more than 30% of the individual donor samples were excluded from all correlation analyses. We set a cut-off of correlation values as biologically meaningful correlations at the Rho values of >= 0.75 for positive/concordant correlations or Rho values of <=-0.75 for negative/discordant correlations. In our gene ontology analysis, we included the markers showing these brain-brain and brain-blood correlations (Fig 1 D-J).

#### Mixed effects analysis & differential gene expression analysis

To better understand the multifactorial dynamics that might influence the relationship or differences between Ant/Ins, sgACC, and blood gene expression patterns, we first conducted a mixed-effects multifactorial analysis of variance using *three* levels of biological regions (Ant-Ins, sgACC, and peripheral whole blood/Paxgene stored blood), and *two* levels of degree of comorbid psychopathology (high number of comorbid psychopathological disorders are reflected in the level of mental disorder complexity vs. low number of comorbid psychopathology. The DESeq2 software (Anders & Huber 2010) was used for the mixed effects analysis with the statistical significance threshold set at p<=0.005.

Our methods to determine high vs. low psychopathology did not selectively include measures of depression per se but rather assessed how many lifetime psychopathological diagnoses and related documented severity of the listed diagnoses per individual donor are captured in our superficially created comparison groups of high (severe/increased psychopathology) > low (severe/absence of psychopathology). To this goal, we used the number of comorbid mental health symptoms and their respective severity in the postmortem diagnoses data to calculate the total of mental health complexity/psychiatric comorbidity and compare high (4 to 7 comorbid diagnosis/symptoms ‘7 being the maximum observed in our sample’) > low (0-3 comorbid diagnosis/symptoms).

We then applied a differential gene expression analysis, using age, sex, and RIN values as covariates, between individuals with a depressive diagnosis (either MDD or DepNOS) versus individuals with any other diagnosis, controlling for both sex and age at death, was assessed using DESeq2 (Anders & Huber 2010). DE analysis was conducted separately in the data sets from each brain region and PAXgene blood at a threshold of p<=0.005. We then examined the depression-related gene expression differences by directly comparing DEGs in the Ant/Ins, sgACC, and blood of depressed versus non-depressed by focusing on brain-brain and brain-blood correlations. We set the significant cut-offs at Log2Foldchange >=1.5 and for those markers meeting this expression threshold to also survive Spearman’s Rho >= +0.25 to +1.0 (concordantly correlated markers) and Spearman’s Rho <= -0.25 to -1.0 (discordantly correlated markers).

As an alternate approach for the identification of markers with consistent patterns of differential expression across region/sample type, we also applied a linear mixed-effects approach (as implemented by the lmerSeq R package) modeling sample type via a random effect and using the same fixed effects structure as was employed in the DESeq2 modeling described above. This approach normalized the RNA-seq data by dividing the mapped read counts by sample-specific size factors estimated using DESeq2 before log2-transformation with a pseudo-count offset of 1.

#### Kolmogorov-Smirnov test

The Kolmogorov–Smirnov (KS) test is a non-parametric method used to determine whether two one-dimensional probability distributions differ (Kolmogorov 1933) (Massey, n.d.). It compares the maximum distance between their cumulative distribution functions (CDFs), making it useful for testing a sample against a reference distribution or comparing two samples. Mathematically, the KS statistics are defined as:

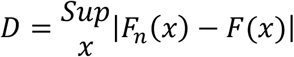

where, *F*_*n*_(*x*) is the empirical CDF of the sample and *F*(*x*) is the reference CDF. The KS test is distribution-free, sensitive to differences in shape and location, and remarkably effective for large sample sizes (Smirnov 1948). Its flexibility makes it valuable in fields ranging from gene expression analysis to quality control, where detecting subtle distributional shifts is essential (Bandyopadhyay et al. 2014). Leveraging this flexibility, Figure 1(A, B, and C) presents a comparative analysis of gene-wise Spearman correlation distributions across Ant/Ins-sgACC and Ant/Ins, sgACC–blood pairings, specifically Ant/Ins–sgACC, Ant/Ins–Blood, and sgACC–Blood. Empirical distributions are visualized as histograms and overlaid with fitted normal distribution curves to reveal patterns in transcriptional co-variation. The Ant/Ins–sgACC distribution demonstrates a righter skew, reflecting a higher prevalence of strong concordant correlations, while the Ant/Ins–Blood and sgACC–Blood distributions are more symmetric and centered near zero, indicating balanced gene-wise correlation patterns with peripheral expression profiles (Drevets, Savitz, and Trimble 2008; Arasappan et al. 2025).

The comparison between Ant/Ins–sgACC and Ant/Ins–blood yields 0.357, Ant/Ins– sgACC and sgACC–blood yielded a KS statistic of 0.422, indicating a significant divergence in their gene-level correlation structures, consistent with region-specific transcriptional coupling or functional specialization within the brain. In contrast, the KS test between Ant/Ins–Blood and sgACC–Blood resulted in a low statistic of 0.070, suggesting minimal distributional divergence and similar patterns of transcriptional association between brain regions and blood transcriptomics. These findings highlight more directionally consistent transcriptomic coordination within Ant/Ins-blood and sgACC-blood regions than between transcriptomics, pointing to shared systemic signatures influencing brain–blood coupling (Jabbi et al. 2020).

#### Pathway Enrichment Analysis

To identify functional pathways associated with gene expression concordance and discordance across brain regions and between brain and blood, we performed pathway enrichment analysis using the *gprofiler2* R package (version 0.2.1) (Raudvere et al., 2019). Genes were grouped based on their Spearman correlation coefficients across tissue comparisons. Specifically, we analyzed gene sets that were concordantly correlated (R >= 0.75) and discordantly correlated (R <= –0.75) between the Ant/Ins and sgACC, as well as between each brain region and blood. For the correlated genes, we subdivided the sets based on the directionality of expression differences (i.e., genes are relatively upregulated in one tissue and downregulated in the other). Enrichment analysis was restricted to the Reactome pathway database to focus on well-curated signaling and molecular pathways relevant to cellular and physiological processes. We applied *gprofiler2*’s gSCS correction method for multiple testing. This approach accounts for the hierarchical structure and gene overlap among functional categories and is specifically designed to reduce false positives in pathway enrichment analyses.

To characterize the molecular signatures of the complex phenotypes understudy using postmortem RNA-seq, we included 14 donors that had relatively high-quality RNA from the two brain regions (subgenual ACC and anterior insula) and whole blood stored in PAXgene tubes. The RNA integrity numbers ranged from 5 to 7.6 for all three samples (sgACC ‘average RIN = 6.68 ±.62’, anterior insula (average RIN = 6.64 ±.63, and whole blood ‘average RIN = 6.68 ±.8’).

#### Brain Dissection

The NIMH Human Brain Collection Core (HBCC) provided the *postmortem* samples for which informed consent is acquired according to NIH IRB guidelines. Clinical characterization, neuropathology screening, and toxicology analyses followed previous protocols (Lipska et al. 2006). The region of interest targeted for dissection of the Ant-Ins was defined as the most anterior portion of the insula encompassing the identified reduced gray matter volume (GMV) in the completed meta-analysis by the authors (Jabbi et al. 2020a). Therefore, the dissected regional volume corresponded to the anterior portion of the Ant/Ins, where the caudate and putamen are approximately equal in size (see Supplementary Figure 1 “**Fig S1A**). Frozen tissue was dissected from the Ant-Ins section for each donor for RNA sequencing. The dissected regional volume from the sgACC was defined as a portion of the ACC Brodmann area 32/25 (**Fig S1B**) (Akula et al. 2021).

#### Brain RNA-Extraction and Sequencing

Twenty-eight total RNA samples isolated from the Ant-Ins and sgACC separately were stored at -80°C for several weeks and subsequently thawed on ice. Each sample was transferred to a PCR-clean Eppendorf low bind twin.tec®, semi-skirted 96-well plate and the final volume was adjusted to 11 µl with nuclease-free dH2O (AM9932), normalized to approximately 500 ng of RNA. RNA was normalized by total rRNA removal using the Ribo-Zero Plus rRNA depletion kit (ref# 20040526). The ribosomal RNA depletion process included three steps according to the manufacturer’s protocol:

Probe Hybridization: Total RNA was hybridized with rRNA depletion probes provided in the kit. Hybridization was performed in a thermal cycler (SimpliAmp by Applied Biosystems) at 95°C for 2 minutes, with a ramp rate of 0.1°C per second, followed by cooling to 37°C.

#### Ribo RNA (rRNA) Depletion

The probe-hybridized total RNA was then treated with an rRNA depletion mastermix (buffer and enzyme provided in the kit) and incubated at 37°C for 15 minutes. Probe Removal: After incubation, kit-provided reagents were added to remove the probes from the depleted RNA, followed by another incubation at 37°C for 15 minutes.

Following globin rRNA depletion, mRNA underwent a 2X bead cleanup using RNA Clean XP beads as recommended. Subsequently, 1 µl of the cleaned rRNA-depleted mRNA was analyzed on a Bioanalyzer to assess the final concentration.

After total rRNA depletion, the resulting mRNA was fragmented and denatured. First-strand and subsequent second-strand syntheses were performed to maintain RNA molecule directionality. The resulting cDNA was purified using a 1.8X Ampure bead cleanup. Next, according to the manufacturer’s recommendation, the purified cDNA underwent A-tailing at its 3’ ends and was ligated with IDT dual index adaptors. Adaptor ligation was stopped using an STL reagent provided in the kit, followed by an additional 0.9X Ampure bead cleanup. Subsequently, the DNA was subjected to 15 cycles of PCR amplification. Finally, after a 1X Ampure bead cleanup, the prepared libraries were ready for downstream processing and sequencing. For the Ant-Ins and sgACC, the total RNA extracted from frozen dissections of the Ant-Ins and sgACC and only samples with RNA integrity numbers (RIN values) greater than 5, as confirmed using the Agilent Bioanalyzer, were used for library preparation. Total RNA was captured using the RiboZero protocol, followed by library preparation, and stranded paired-end sequencing was performed on the RNA samples using the Illumina HiSeq 2500 system. We obtained an average of one hundred and forty million reads per sample, totaling ∼54 billion reads. After quality control, reads were mapped to human genome build 38 using Hisat2 (Pertea et al. 2016). Finally, gene and transcript counts were obtained using StringTie (Pertea et al. 2016).

For both Ant-Ins and sgACC, any genes expressing 0 in 80% or more samples were filtered out to remove low-count genes from further analysis. Next, the abundances were normalized using DESeq2 and transformed with variance stabilizing transformation (a transformation to yield counts that are approximately homoscedastic, having a constant variance regardless of the mean expression value). Finally, Principal Component Analysis was performed using 25% of the highest variance genes to explore the underlying data’s structure and the most significant sources of variance. Lastly, genes with an expression value of 0 in 80% of samples or more were removed from further analysis to correct for sporadically large fold-change outliers.

#### Blood RNA-Extraction

Blood was collected by the medical examiner in PAXgene Tubes (2.5 mL, BD Biosciences Cat. # 762165). They were maintained at room temperature for 2-72 hours to ensure complete lysis of blood cells. A step-down freezing method was then performed by freezing the tubes at -20 C for 24 hours, followed by transfer to a -80 C freezer. The tubes were then equilibrated at room temperature for 2 hours before starting RNA extraction. RNA extraction followed the manufacturer’s instructions for the PreAnalytix PAXgene Blood RNA Kit (Cat#762164). Specifically, the PAXgene tube samples are removed from the freezer and left to defreeze, and then 350 µL Buffer BR3 is pipetted into the PAXgene RNA spin column. The sample is then centrifuged for 1 minute at 8000–20,000 × g, and the spin column is placed in a new 2 mL processing tube and discard the old processing tube containing flow-through. A 10 µL DNase I stock solution was added to 70 µL Buffer RDD in a 1.5 mL microcentrifuge tube. The sample is mixed by gently flicking the tube and centrifuging briefly to collect residual liquid from the sides of the tube. If processing, for example, 10 samples, add 100 µL DNase I stock solution to 700 µL Buffer RDD. Use the 1.5 mL microcentrifuge tubes supplied with the kit. The DNase I incubation mix (80 µL) is pipetted directly onto the PAXgene RNA spin column membrane and placed on the benchtop (20–30°C) for 15 minutes. After DNAse treatment, the purified RNA from whole blood was used separately per individual to determine RNA quality as measured in RNA integrity number (RIN) values using Agilent 6000 RNA Nano Kit consisting of the microfluidic chips, Agilent 6000 RNA Nano ladder, and reagents on Agilent 2100 Bioanalyzer. Only blood RNA samples with RIN values >=5 were included in the study. RNA concentration was measured with the NanoDrop One/Onec Spectrophotometer (Thermo Scientific Part Number: ND-ONE-W), and RNA quality was assessed with the Agilent Bioanalyzer 2100, using the Agilent RNA 6000 Nano Kit (Agilent Technologies, Cat. # 5067-1511), as per the manufacturer’s specification.

#### Blood Globin RNA Depletion and RNA-sequencing

Twenty-one total RNA samples isolated from Human blood containing the Paxgene were stored at -80°C for several weeks and subsequently thawed on ice. The total RNA was transferred to a clean PCR Eppendorf low-bind twin.tec®, semi-skirted 96 well plate, and the final volume was adjusted to 10 µl with nuclease-free dH2O (AM9932) and normalized to approximately 500 ng. This normalized RNA underwent Globin rRNA removal using the Illumina TruSeq Stranded Total RNA Lib Prep Globin Kit (ref# 20020612).

An equivalent volume (v/v) of rRNA binding buffer and Globin RNA removal mix (provided in the kit) was added to the total RNA, and the mixture was incubated at 68°C for 5 minutes in a thermal cycler (SimpliAmp by Applied Biosystems). Subsequently, an appropriate amount of rRNA removal beads (provided in the kit) was added to the RNA and Globin removal reagent mix. After a brief incubation of 1 minute on a magnetic stand (Alpaqua FLX), the entire mixture, excluding the beads, was transferred to a new plate. As recommended, the globin rRNA-depleted mRNA underwent a 1.8X bead cleanup using RNA Clean XP beads. Finally, 1 µl of the cleaned rRNA-depleted mRNA was analyzed on a Bioanalyzer to determine the final concentration.

Following the depletion of globin mRNA, the resultant mRNA was fragmented and primed for the First strand synthesis. Subsequently, first-strand synthesis followed by second-strand synthesis was performed to maintain RNA molecule directionality. The resulting cDNA underwent purification using a 1.8X Ampure bead cleanup. Next, A-tailing was carried out at the 3’ ends of the purified cDNA, followed by ligation with Illumina dual index adaptors (GSAF lab) at a concentration of 1 µM. The adaptor ligation was terminated using an STL reagent provided in the kit, and the index-ligated DNA underwent an additional 1X Ampure bead cleanup. The DNA was then subjected to 15 cycles of PCR amplification. Finally, after a 1X Ampure bead cleanup, the prepared libraries were ready for downstream processing and sequencing.

## Data Availability

RNA sequencing Data, and demographics data.

## Acknowledgments

The NIMH Human Brain Collection Core (HBCC) provided RNA samples for all the donors included in this study, and we thank Drs. Stefano Marenco, Pavan Auluck, and HBCC colleagues thank you for providing the samples and related metadata. The HBCC is supported by the NIMH-IRP project ZIC MH002903. We thank Jessica Podnar and the University of Texas genome sequencing and facility (GSAF) colleagues for RNA-seq support. This work was supported by the KL2TR002646 (with Dr. Joel Tsevat et al. as PI/co-Is) grant from NCATS and by the Dell Medical School Mulva Clinics for the Neurosciences, UT Austin, and NIH 1R01MH134791-01.

## Author Contributions

DWylie, DArasappan, PRathouz, and MJabbi designed the study. DWylie, DArasappan, UKhatri, MDwortz, and MJabbi performed the analyses. MJabbi drafted, and all other authors contributed to writing the manuscript.

## SUPPLEMENTARY MATERIALS

### StripCharts

The strip_chart plot shows the genes analyzed from that particular gene set. The y-axis in the strip charts is the difference in expression levels between the blood sample for each individual and the average of the two brain regions for the same individual, grouped by morbidity group, as this is similar to what the interaction term analysis is using as input to the enrichment analysis for the GO terms.

### Pathways and GO RESULTS Explanation of Brain-Blood Transcriptomics Main

#### Effects and Interaction

The volcano_plot summarizes the gene set analysis for enrichment in genes with more significant Morbidity: Region=Blood interaction terms, while the response_to_cytokine. A bit more regarding the volcano plot: the x-axis is mapped to a metric known as the “**A**rea **U**nder the **<u>R</u>**<u>eceiver **O**perating **C**haracteristic</u> curve,” or AUROC (often referred to as AUC). This can provide a useful effect size measure for gene set enrichment analysis as it has a valid interpretation in this context: The AUROC for gene set *G* measures the likelihood that a randomly chosen gene *g* selected from the gene set will have a larger value of the differential expression statistic than will a randomly chosen gene *h* selected from all genes **not** included in the gene set *G*.

Thus, AUROC values close to 0.5 indicate no enrichment in differentially expressed (DE) genes, while AUROC values much less than 0.5 indicate enrichment in discordant DE genes, and AUROC values much more significant than 0.5 indicate enrichment in concordant DE genes. (Note that AUROC must be between 0 and 1 in all cases. The y-axis in the volcano plot is just the (negative) log-scaled p-values from the GOMWU enrichment testing method for the GO term gene sets. Those gene sets for which the false-discovery rate (FDR)-adjusted p-values are less than 0.01 are colored in red (0.01 is a conservative threshold here, so anything colored red certainly shows strong evidence of at least some degree of enrichment in DE genes). The points’ size in the plot indicates the number of genes included—larger points represent larger gene sets.

**Supplementary Figure 1.**
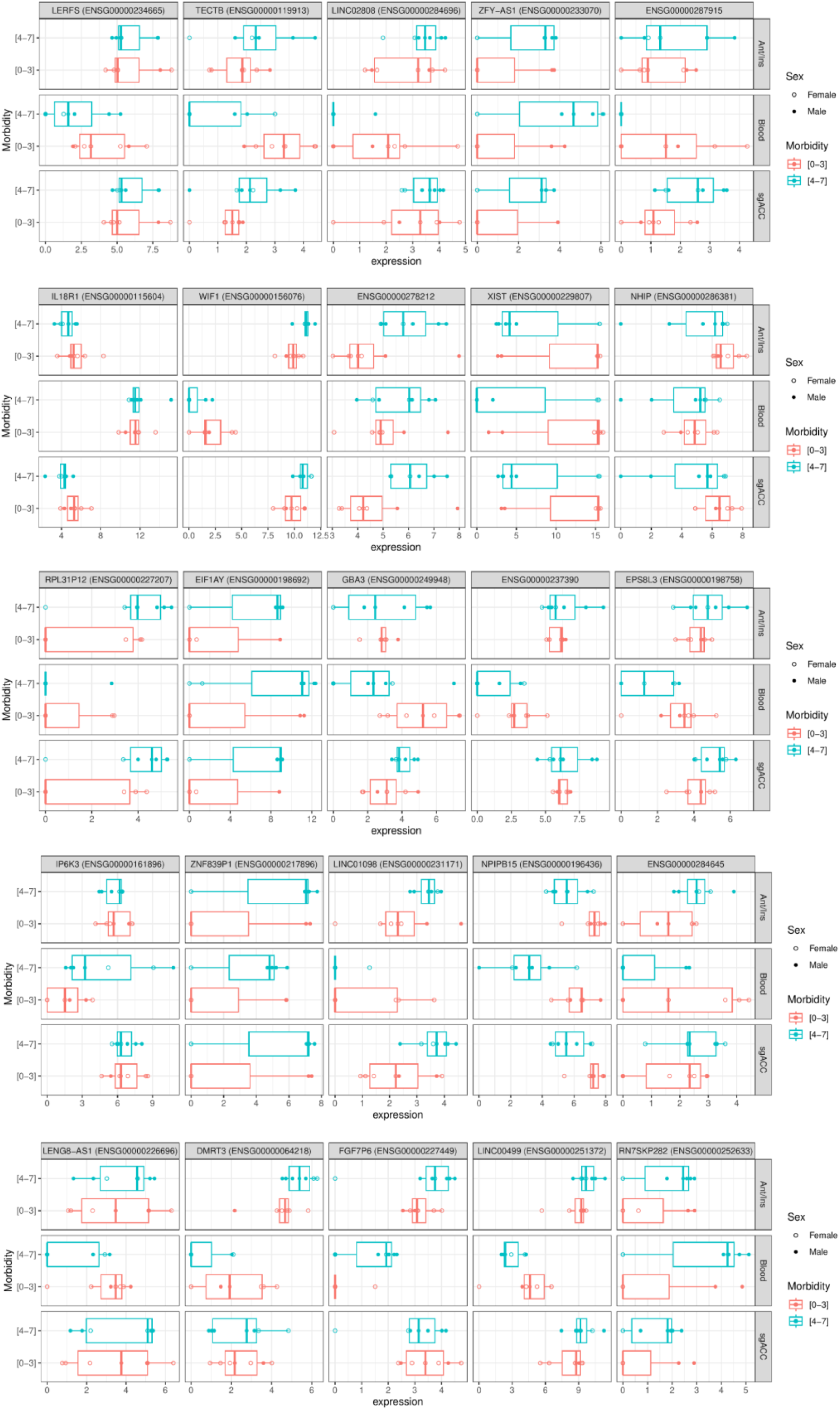
Expression levels (after DESeq2 normalization and log2 transformation using an additive pseudocount of 1, indicated on the x-axis of each panel) for the top 25 genes by mixed-effects interaction analysis (interaction term for sample type (brain region or blood) x morbidity group (morbidity 0-3 vs. 4-7)).

We applied GO-MWU using the Gene Ontology Mann-Whitney U testing. We outlined in the following online resources: https://github.com/z0on/GO_MWU to identify Gene Ontology (GO) gene sets whose constituents were enriched in more extreme mixed-effects test statistic values relative to the background distribution of such differential expression statistics. The GO-MWU method analyzes each GO category G within specified size limits (minimum of 10 observed genes, maximum of 50% of all observed genes) and tests whether the scores associated with genes within G differ significantly from the scores associated with genes not within G using the non-parametric Mann-Whitney U test.

**Supplementary Figure 2.**
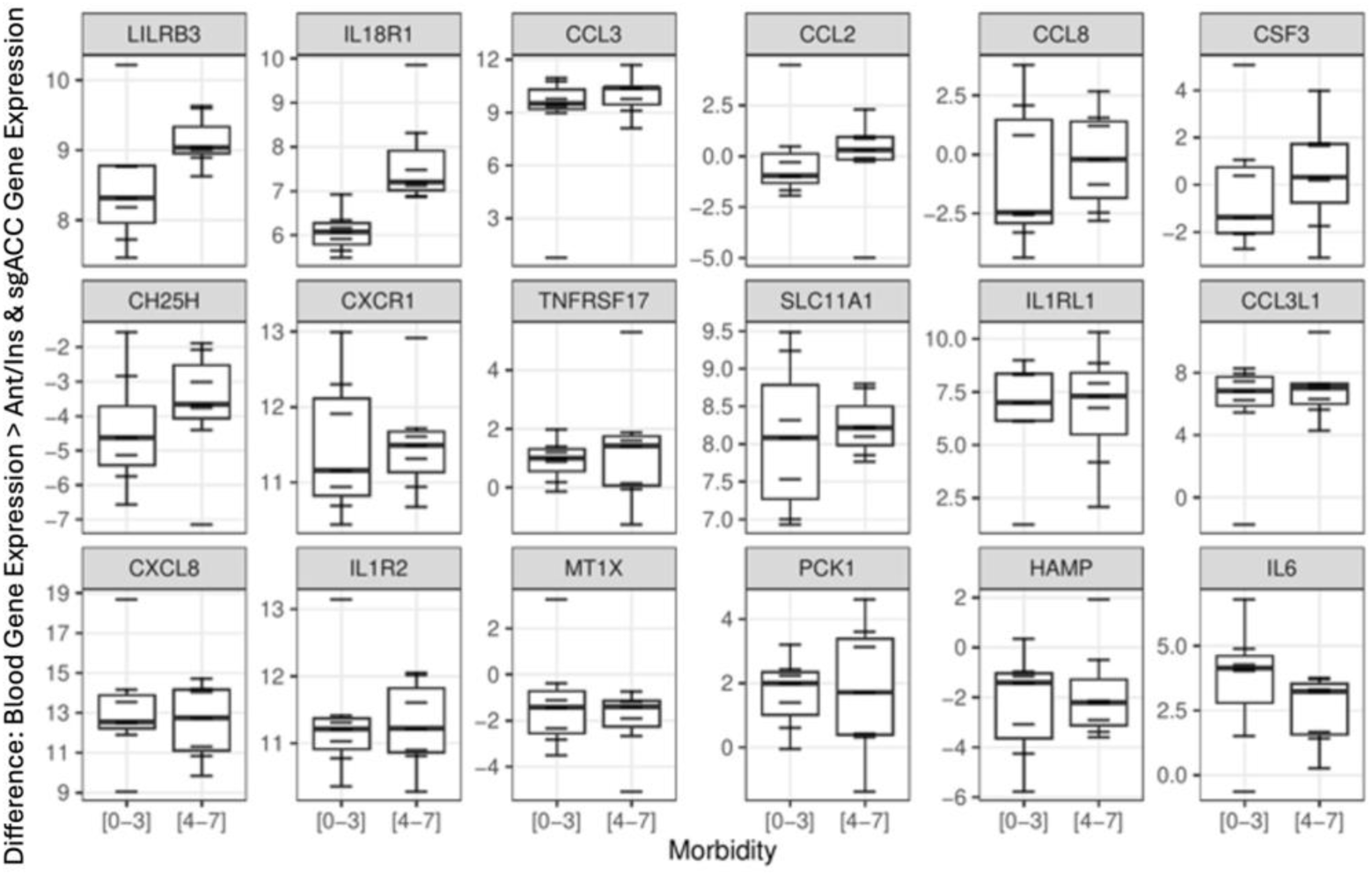
Box-and-whisker plots of within-individual difference in expression levels (blood minus the average of the two brain regions, all calculated after DESeq2 normalization and log2 transformation using an additive pseudo-count of 1; indicated on the y-axis of each panel) for the indicated cytokine genes. Expression levels are separated (on the x-axis) within each panel by morbidity group (morbidity 0-3 vs. 4-7).

## SUPPLEMENTARY METHODS

### RNA-Extraction of Ant-Ins & sgACC

The HBCC further pulverized all dissected tissues separately and aliquoted 50mg from each sample for standardized total RNA processing. Specifically, RNeasy Lipid Tissue Mini Kit (50) was used for RNA purification using the 50 RNeasy Mini Spin Columns, Collection Tubes (1.5 ml and 2 ml), QIAzol Lysis Reagent, RNase-free Reagents, and Buffers kit from Qiagen. DNase treatment was applied to the purified RNA using a Qiagen RNase-Free DNase Set (50) kit consisting of 1500 Kunitz unit’s RNase-free DNase I, RNase-free Buffer RDD, and RNase-free water for 50 RNA minipreps. After DNAse treatment, the purified RNA from pulverized AIAC was used separately per individual to determine RNA quality as measured in RNA integrity number (RIN) values using Agilent 6000 RNA Nano Kit consisting of the microfluidic chips, Agilent 6000 RNA Nano ladder, and reagents on Agilent 2100 Bioanalyzer. Samples with RIN < 6 were excluded from the study.

### Blood Processing

Blood was collected by the medical examiner in PAXgene Tubes (2.5 mL, BD Biosciences Cat. # 762165). They were maintained at room temperature for 2-72 hours to ensure complete lysis of blood cells. A step-down freezing method was then performed by freezing the tubes at -20 C for 24 hours, followed by transfer to a -80 C freezer. The tubes were then equilibrated at room temperature for 2 hours before starting RNA extraction. RNA extraction followed the manufacturer’s instructions for the PreAnalytix PAXgene Blood RNA Kit (Cat#762164). RNA concentration was measured with the NanoDrop One/Onec Spectrophotometer (Thermo Scientific Part Number: ND-ONE-W) per the manufacturer’s specifications. We then assessed RNA quality using the Agilent Bioanalyzer 2100 using the Agilent RNA 6000 Nano Kit (Agilent Technologies, Cat. # 5067-1511), per the manufacturer’s specifications.

**Supplementary Figure 3.**
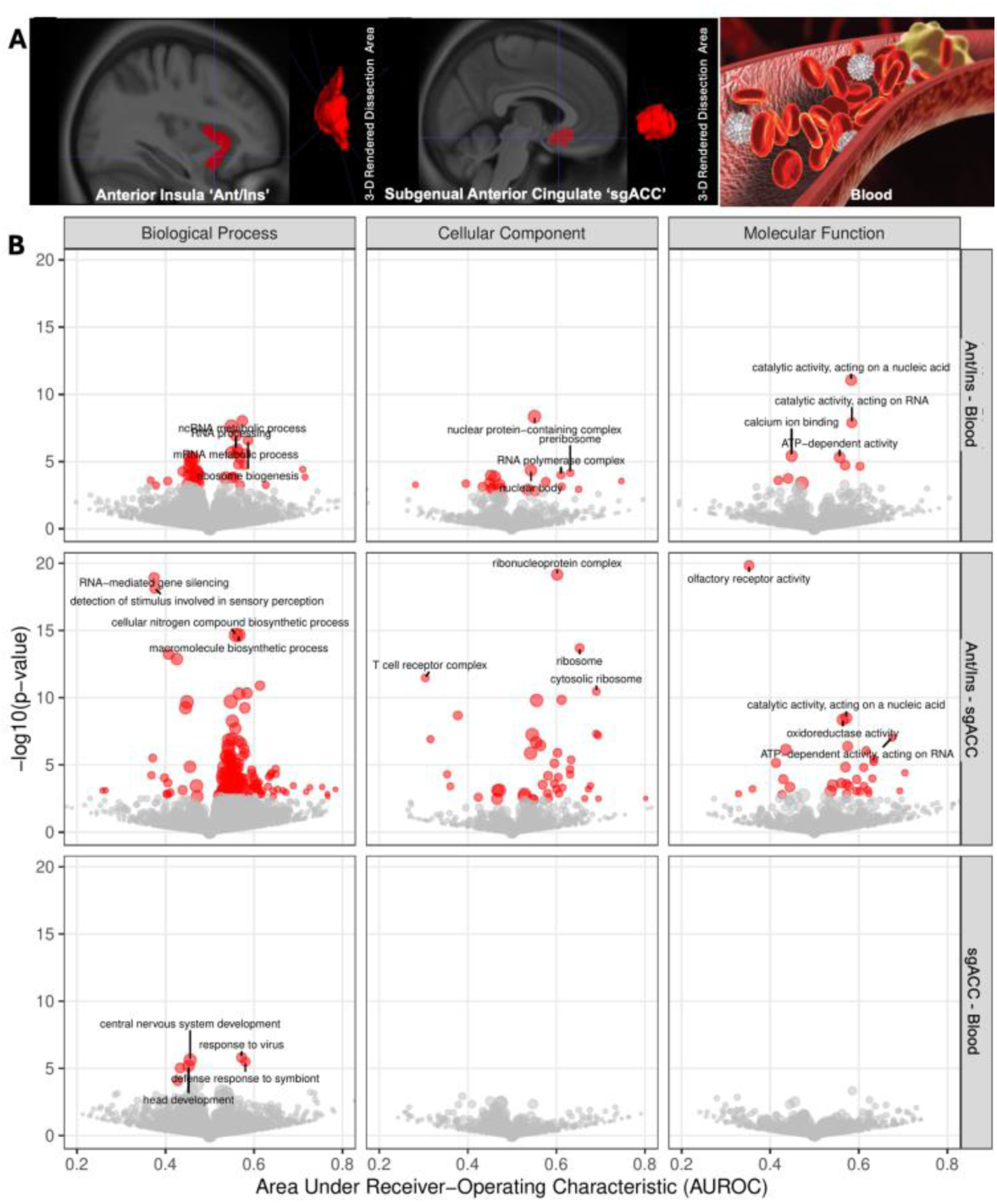
Volcano-style plots presenting evidence of statistical significance (log-transformed p-values, y-axis) compared to effect size estimates (Area Under Receiving Operating Characteristic, or AUROC, scores, x-axis; a value greater than 0.5 indicates over-expression predominates, while AUROC values less than 0.5 indicate predominance of under-expression) for GO-MWU gene set analyses. Each point corresponds to a single analyzed gene set; points are colored red if their p-value is low enough that they would be declared significant using a Benjamini-Hochberg FDR threshold of 0.1 (10%). The four gene sets with the lowest p-value in each panel are labeled if they met the FDR cutoff.

